# Effectiveness of Community-Based Rehabilitation (CBR) centres for improving physical fitness for community-dwelling older adults: a systematic review protocol

**DOI:** 10.1101/2022.03.13.22272309

**Authors:** Wei Xin, Dan Xu, Zulin Dou, Angela Jacques, Josephine Umbella, Anne-Marie Hill

## Abstract

**Introduction:** The increasing ageing population has become a substantial challenge for both health care and social services in many Asian countries. There is a high incidence of chronic diseases and comorbidities in older populations, leading to impairments and functional disability. Functional disability may result in loss of independence, reduced quality of life and increased care needs. Community-based rehabilitation (CBR) aims to promote equality of opportunity and improve the social inclusion of individuals living with disability. CBR also provides rehabilitation to improve physical, mental, and social outcomes. However, there is limited evidence regarding the effectiveness of CBR for improving older adults’ physical fitness. The aim of this systematic review is to synthesise the evidence for the effectiveness of interventions delivered by CBR centres on physical fitness of community-dwelling older adults in Asian countries.

**Methods and analysis:** A search on four English databases (CINAHL, Medline, Scopus and Proquest) and two Chinese databases (China National Knowledge Internet and Wanfang Database) will be conducted, from inception to 15 November 2021. Both English and Chinese publications will be included. Experimental and quasi-experimental studies using any type of control group will be included. The primary outcomes are physical fitness (capacity to perform activities and tasks). Secondary outcomes are performance of activities of daily living and health-related quality of life. The quality of all included studies will be assessed using the Joanna Briggs Institute (JBI) standardised critical appraisal tools. Two reviewers will independently complete study screening, selection, quality appraisal, and data extraction. Quantitative data where possible will be pooled in statistical meta-analysis. All statistical analyses will be performed using Review Manager (Rev Man) V.5.3 software.

**Ethics and dissemination:** Ethical approval is not required for this review. The findings of this review will be disseminated electronically through a peer-reviewed publication and conference presentations.

**PROSPERO registration number:** CRD42021292088

**Strengths and limitations:**

- Findings and evidence in this review will be summarised and graded using the Grading of Recommendations, Assessment, Development and Evaluation Pro (GRADEPro) approach.
- A comprehensive literature search using both English and Chinese language databases will be conducted.
- Studies included in the review may measure different outcomes which may limit pooling in meta-analysis.
- Differences in populations and interventions delivered in the included studies may result in high levels of heterogeneity, leading to less certainty about the recommendations from the review.

## Background

Population ageing has become a major public health challenge worldwide, although this is occurring at varying speeds in different counties.^1^ According to the United Nations, a country with a proportion of older adults (aged 60 years or older) comprising of more than 10% of the total population, is identified as an ageing country.^2^ Asia is the largest earth continent with 60% of the world population.^3^ Most Asian counties are facing unprecedented growth of the ageing population and have stepped into an ageing society.^4^ The United Nations Economic and Social Commission for Asia and the Pacific (ESCAP) estimates that the proportion of the older adult population living in Asian countries will double from 12.4% in 2018 to reach more than 25% (1.3 billion) by 2050.^3^ In China the older adult population was over 264 million in 2016, accounting for more than 18% of the total population. It is estimated that older adults in China will account for 30% of the total population (over 400 million) by 2050.^5^ This significant demographic change will have considerable impacts on social services, health, economic and social care, presenting both challenges and opportunities.^4^

The prevalence of chronic diseases among older adult populations is increasing worldwide due to both non-modifiable risk factors (such as ageing processes and genetics) and modifiable risk factors (such as smoking, dietary, and exercise patterns).^6^ An epidemiological study investigating chronic diseases in China found that the top three chronic diseases are hypertension, diabetes and hypercholesterolemia, and their prevalence rates are 58.3%, 19.4% and 10.5% respectively.^7^ These chronic diseases contribute to increases in the incidence of cardiovascular, neurodegenerative and metabolic diseases, thereby leading to increased functional disability in the ageing population.^8^ Functional disability is predictive of reduced walking, talking, and bathing abilities, and higher incidence of falls, social isolation, and dependency, resulting in reduced ability to participate in the community and maintain independence in ADL.^9^

Long-term care needs required for functional disability will increase due to the increasing ageing population and are predicted to be the primary determinant of the rising burden to the healthcare system.^10^ It is estimated that, in the following one and half decades, healthcare costs for older adults will account for about 65% of the Chinese government health burden and 44% of the Indian government health burden.^10^ WHO defines functional ability as “having the capabilities that enable all people to be and do what they have reason to value”, including abilities to meet their basic needs, learn, grow, and make decisions, be mobile, build and maintain relationships, and contribute to society.^11^ This includes performance of basic activities of self-care, such as dressing, walking, or eating, which are referred to as activities of daily living (ADL).^12^ Physical fitness is a core component of functional ability and is defined as the ability to carry out daily tasks and perform physical activities in a highly functional state, often as a result of physical conditioning.^13^ Physical activity is defined as any bodily movement produced by skeletal muscles that requires energy expenditure and refers to all movement including during leisure time, transport to and from places or as part of a person’s work.^14^

There is strong evidence that a range of rehabilitation interventions are effective in improving physical fitness and ADL, which in turn improves or maintains quality of life for community-dwelling ageing populations.^15 16^ Systematic reviews and meta-analysis demonstrate significant reduction in fall rates and improvements in dynamic balance, static balance, functional performance, and quality of life in older adults who received physical exercise, progressive resistance strength training (PRT) exercises, Tai-Chi, and dancing compared to controls.^17-20^ Reviews have also found that ADL, such as bathing and dressing, are improved by strength, endurance, balance, and functional training.^21-24^ However, rehabilitation interventions are not yet widely available in Chinese communities, as there is limited ability for community centres to provide rehabilitation treatment, health professional support and resources.^25^ A Chinese study published in 2021 reported that while 11.1% of older adults required rehabilitation services, less than 1% of older adults actually received various rehabilitation services in their community.^26^ These findings highlight the limited availability of community rehabilitation services for older adults in China and the need to address this service gap to reduce the growing health burden due to functional disability.

Community-based rehabilitation (CBR) was originally promoted by the World Health Organization in 1978, and further defined in 2004 as a key strategy that aims to meet the rehabilitation needs, equalization of opportunities and social inclusion of people living with disabilities in the general community.^27^ CBR delivers a broad range of health services including rehabilitation and physical exercise training within the community, supervised by health professionals but using predominantly local resources.^28^ Compared with existing health care systems, the CBR system has the potential to provide health care services for the community in a timely and efficient manner without cumbersome and compartmentalized local administration. This promotes strong and convenient links between older adults living with disabilities and their local health-care system.^29^ A study conducted in Nepal that investigated the cost-effectiveness of CBR programs reported that the programs averted 1065 disability-adjusted life years from 2013 to 2015.^30^

However, there is limited evidence about the effectiveness of CBR for improving physical fitness or functional ability of older adults.^28 31 32^ CBR has been found to have some limitations, which have been summarised as not being customised to the local settings, absence of guidelines or frameworks for commencement and implementation, shortages of resources, lack of professional health care workers and multidisciplinary teams to deliver rehabilitation services.^29^ Previous systematic reviews have provided limited evidence for the effectiveness of CBR for improving health outcomes for community-dwelling older adults. A systematic review that included 15 studies found that CBR is effective in improving physical functional disabilities (such as performance of ADL and walking ability), mental functional disability and quality of life for people with living with disabilities (including people with stroke, arthritis, chronic obstructive pulmonary diseases, and mental disabilities) living in low- and middle-income countries. However, this review was not limited to any particular age group and did not undertake sub-group analyses based on age. An integrative review that evaluated the effectiveness of CBR on self-care management for older adults, found that CBR can improve health behaviours and other outcomes, however, this review did not include outcomes that related to physical fitness or ADL.^33^

A preliminary literature search conducted through Google Scholar, PubMed and CINAHL found no systematic reviews that are undergoing or have been published evaluating the effectiveness of interventions delivered in the CBR centres on physical fitness or ADL for community-dwelling older adults in Asian countries. There is a gap in evidence about whether CBR delivered interventions can improve older adults’ performance of ADL or physical fitness as well as other important health outcomes, such as health related quality of life. This systematic review will summarise the best available evidence for CBR interventions that aim to improve ADL or physical fitness for older adults, living in Asian countries. There is a rapidly increasing number of countries in Asia with ageing populations.^1^ The focus on countries with similar cultures and histories will assist in providing high quality evidence for Asian health systems as well as health systems in other counties that use CBR. The sharp increase in the ageing population and the resource constraints of health systems mean it is important to synthesise the best evidence for whether CBR interventions are effective in improving older adult’s physical fitness and performance of ADL.

### Review objective

The objective of the review is to synthesise the best available evidence for the effectiveness of interventions performed in CBR centres on i) physical fitness ii) activities of daily living and health related quality of life; for older adults aged over 60 living in Asian countries.

## Methods

The review protocol has been registered on Prospero (ID: CRD42021292088). The results of the review will be reported according to the Preferred Reporting Items for Systematic Reviews and Meta-Analyses (PRISMA) 2020 Guidelines.^33^

### Inclusion criteria

#### Types of participants

This review will consider studies that include community-dwelling adults aged 60 years or over living in Asian countries. The MeSH term “Asia” has five parts, including Central, Northern, South-Eastern, Western, and Far East. China is included in the East Asia region. There are 48 countries and regions in Asia. Studies that enrol participants less than 60 years old but where the mean age of the group is over 60 years old will also be considered for inclusion this review. Studies that enrol older adults receiving palliative care will be excluded.

#### Types of settings

Studies that are conducted by centres that provide CBR will be eligible for inclusion. CBR centres provide services that include physiotherapy, exercise training, exercise, education, and medical services. Services provided by hospitals, including outpatient services, individual community medical practitioners, and home visiting nurses, will be excluded.

#### Types of intervention(s)/phenomena of interest

Primary research evaluating the effectiveness of interventions delivered by CBR centres, either single or multiple component interventions, will be considered for inclusion in this review. Studies evaluating interventions conducted by CBR centres that will be eligible for inclusion will include, but are not limited to, those that deliver multidisciplinary interventions, physiotherapy, occupational therapy (including one-one or group interventions), exercise training (including aerobic, strength, dance, or Tai Chi training), functional training or health education (including one-one/group/written/materials). These interventions must be provided by CBR centres. Studies that evaluate interventions provided in the home will be included if the participants are assessed through the CBR centre and the program is conducted as an outreach of the CBR centre.

#### Comparator(s)/control

This review will only include primary studies that clearly state that the only difference between groups is the use of an intervention delivered by the CBR centre.

Comparators can be usual care or another treatment, such as a home program or health intervention, health education or medications.

#### Types of outcomes

The primary outcomes will be physical fitness (capacity to perform activities and tasks), measured by tools assessing physical function, including, but not limited to, balance, strength, functional mobility, walking, physical activity, and endurance. Measurement tools may include, but are not limited to, BERG balance scale,^34^ Timed up and go test,^35^ gait speed or the 6-minute walk test.^36^ Only studies that measure physical outcomes with validated assessment tools will be included. Cognitive, emotional, and social functional outcomes will be excluded. Studies that measure blood markers such as cholesterol or blood glucose levels will be excluded unless they also include data for outcomes measuring functional performance that can be extracted. Secondary outcomes are performance of ADL measured by tools including but not limited to the Katz tool,^37^ the Lawton instrumental activities of daily living (IADL) scale;^38^ and Health-related quality of life measured by tools including, but not limited to, the EQ5D5L.^39^

#### Types of studies

Experimental and quasi-experimental study designs, including randomised controlled trials (RCTs), or quasi-experimental studies will all be considered for inclusion in this review. Trials that use any type of control group will be included. Case control studies, observational cohort studies or qualitative studies will be excluded. Mixed methods studies will be considered if they contain an experimental phase and report quantitative data that measure an intervention of interest with a comparator.

#### Types of languages

Both English and Chinese publications will be included in this systematic review.

#### Search strategy

This review search strategy aims to ensure the literature search is as wide and thorough as possible so that all relevant published studies can be found and synthesised in the analysis. The search will follow the three-step strategy described by the Joanna Briggs Institute manual including both electronic and manual searches.^40^ Initially, a preliminary web search, including Google Scholar and PubMed, will be conducted to find existing, similar systematic reviews, assessing the volume of potentially relevant studies, and identifying index terms and keywords. Preliminary keywords will be identified and discussed with a senior health librarian at Curtin University. Secondly, all identified keywords and index terms will be searched across the following databases for relevant studies to complete a comprehensive, systematic literature search. Four English electronic databases will be searched: CINAHL, Medline, Scopus and Proquest. Two Chinese electronic databases will be searched: China National Knowledge Internet and Wanfang Database. Grey literature will be searched in OpenGrey. All searches will be reported in the results as recommended in the PRISMA guidelines.^41^ An example of a search strategy is presented in Table 1. Thirdly, the reference list of all identified publications will be hand-searched for additional studies not previously identified during the first or second step of the search. Titles and abstracts of all studies will be screened by two independent reviewers who will examine the studies to determine if they meet the inclusion criteria. Any disagreements between the two reviewers will be resolved by consulting a third independent reviewer until a consensus is reached.

**Table 1.** Literature search strategy for CINAHL

|  |  |
| --- | --- |
| S1 | MH "Aged+" OR (elder* or aged of "old* adults*" or geriatric) |
| S2 | MH "Asia+" OR (china or indonesia or pakistan or bangladesh or japan or philippines or vietnam or turkey or iran or thailand or myanmar or korea or iraq or afghanistan or "saudi arabia" or uzbekistan or malaysia or yemen or nepal or "sri lanka" or kazakhstan or syria or cambodia or jordan or azerbaijan or "united arab emirates" or tajikistan or israel or laos or lebanon or kyrgyzstan or turkmenistan or singapore or oman or palestine or kuwait or georgia or mongolia or armenia or qatar or bahrain or "timor-leste" or cyprus or bhutan or maldives or brunei or taiwan or "hong kong" or macao or macau) |
| S3 | (Communit* N8 (rehab* or therap* or physio*) OR MH<br>"Rehabilitation, Community-Based" |
| S4 | TI communit* OR AB communit* |
| S5 | MH "Rehabilitation" or "Activities of Daily Living+" or<br>"Recreational Therapy" or "Occupational Therapy" or "Physical Therapy+" or "Dance Therapy" or "Rehabilitation, Cardiac" or<br>"Rehabilitation, Pulmonary" or "Chest Physical Therapy" or<br>"Rehabilitation, Geriatric" or "Sensory Motor Integration" or "Early Ambulation" or "Rehabilitation Centers" |
| S6 | S4 AND S5 |
| S7 | TI (rehab* or therap* or physio*) OR AB (rehab* or therap* or physio*) |
| S8 | "Community Health Services" or "Community Health Centers" or |
|  | "Senior Centers" or "Health Services for the Aged" |
| S9 | S7 AND S8 |
| S10 | S3 OR S6 OR S9 |
| S11 | S1 AND S2 AND S10 |

#### Assessment of methodological quality

All eligible studies will be assessed for methodological quality, including risk of bias of individual studies, by two independent reviewers (two for Chinese and two for English articles) before inclusion in the review using the Joanna Briggs Institute standardized critical appraisal tools (JBI https://jbi.global/critical-appraisal-tools) for RCTs and quasi-experimental trials. Any disagreements between the two reviewers will be discussed with a third reviewer to reach consensus. Study selection and inclusion will be presented in a PRISMA flow chart (see Appendix I). Studies excluded after full text review will be recorded and reasons for exclusion provided.

#### Data extraction

Quantitative data will be extracted from all included studies by two reviewers (two for Chinese and two for English articles) using the standardised data extraction tool from the JBI reviewers’ manual.^40^ Detailed information of the included studies extracted will include publication date, authors’ names, types of interventions, populations, study methods, and data measuring outcomes relevant to the review. Missing data from any of the included studies will be sought from the authors. If sufficient data are not obtained from authors studies will be narratively reported and implications of any missing data will be discussed.

#### Data synthesis

Quantitative data from two or more studies will be pooled, if possible, in statistical meta-analysis using Review Manager 5.^42^ All results will be subject to double data entry. Effect sizes will be expressed either as weighted (or standardised) mean differences (for continuous data) or odds ratios (for categorical data), with 95% confidence intervals calculated for analysis, depending on the outcome measures used in the studies. Standard Chi-squared and I-squared tests will be used to assess heterogeneity.^43^ The choice of statistical model (random or fixed effects) for meta-analysis will be based on the criteria outlined by JBI guidelines.^44^ Data that cannot be pooled will be reported using narrative synthesis.^45^ Grading of Recommendations, Assessment, Development and Evaluation Pro (GRADEPro) will be utilised to create the GRADE approach for grading the certainty of evidence and a summary of findings.^46^ The GRADE approach also rates the risk of bias at the outcome level.

## Data Availability

This is a protocol for a systematic review without any data.

## Author contributions

Wei Xin was primarily responsible for drafting the systematic review protocol with support from Anne-Marie Hill and Dan Xu. Wei Xin, Anne-Marie Hill, Dan Xu and Zulin Dou were responsible for study design. Angela Jacques contributes to the design and the statistical analysis plan, Josephine Umbella contributes to search strategy design, data extraction and critical appraisal. All authors critically reviewed the systematic review protocol for its content and approved the final version of the systematic review protocol for submission.

## APPENDIX I PRISMA flow chart

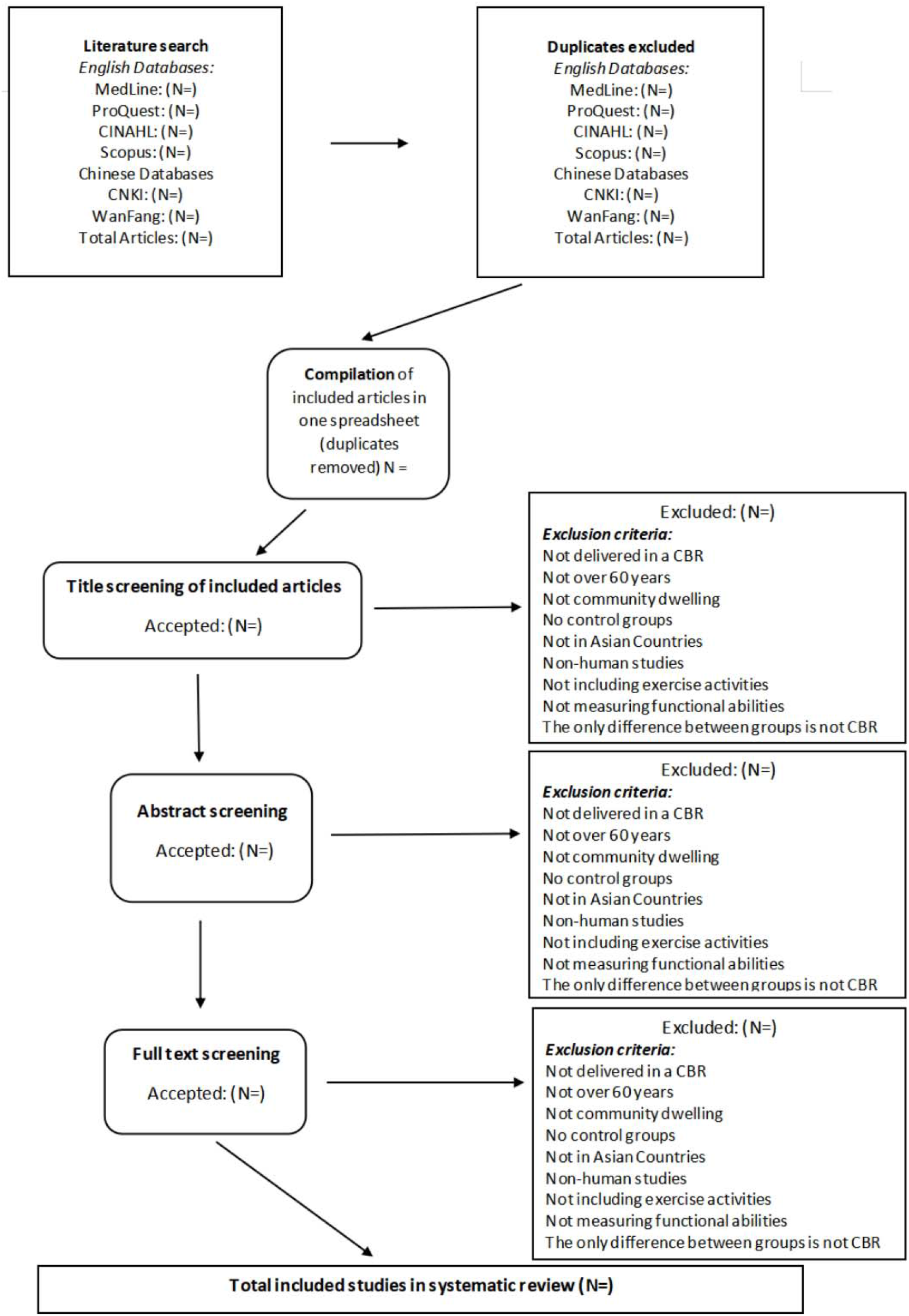

